# Natural Language Word-Embeddings as a glimpse into healthcare at the End Of Life

**DOI:** 10.1101/2021.07.22.21260874

**Authors:** Shun Lau, Zeljko Kraljevic, Mohammad Al-Agil, Shelley Charing, Alan Quarterman, Harold Parkes, Victoria Metaxa, Katherine Sleeman, Wei Gao, Richard Dobson, James T Teo, Phil Hopkins

## Abstract

**Introduction:** Planning in advance and personalised discussions on limitation of life sustaining treatment (LST) is an indicator of good care. However, there are many linguistic nuances and misunderstandings around dying in hospital as well as inaccuracy in individual-level prognostication.

**Methods:** Using unsupervised natural language processing (NLP), we explored real-world terminology using phrase clusters with most similar sematic embeddings to “Ceiling of Treatment” and their prognostication value in the electronic health record of an urban teaching hospital.

**Results:** Word embeddings with most similar to “Ceiling of Treatment” clustered around phrases describing end-of-life care, ceiling of care and resuscitation discussions. The phrases have differing prognostic profile with the highest 7-day mortality in the phrases most implicitly referring to end of life -“terminal care”, “end of life care” (57.5%) and “unsurvivable” (57.6%).

**Conclusion:** NLP can quantify and analyse real-world end of life discussions around prognosis and appropriate LST.

**Patient-friendly Summary:** (by expert patients: Sherry Charing, Alan Quarterman, Harold Parkes)

Discussions between doctors, patients and family in deciding what is the appropriate maximum treatment a specific patient should have based on their clinical condition is complex. Discussions, often involving expressions regarding “End Of Life” care are used to describe the maximum invasive treatments a patient should have or would want. There are a range of expressions used, many with overlapping meanings which can be confusing, not only for the patient and family, but also for doctors reading the patient’s clinical notes. In this study, a computational approach using Artificial Intelligence to read clinical patient notes was carried out by looking at thousands of patient records from a large urban hospital. Expressions that doctors use to describe these discussions were analysed to show the associations of particular words and phrases in relation to mortality. Using a computer analysis for this study it was possible to quantify the use of these expressions and their relation to the “End Of Life”. Through this AI-based approach, real-world use of phrases and language relating “End Of Life” can be analysed to understand how doctors and patients are communicating, and about any possible misunderstandings of language.

## Introduction

Planning in advance for ‘End Of Life’ care is a complex and sensitive area of healthcare, and there is significant room for misunderstandings^1–3^. However, such discussions and advance decisions can be mishandled without personalised counselling as misperceptions may arise about what kinds of treatments are referred to^4^. Phrases such as ‘ceiling of treatment’ and ‘treatment escalation plans’ attempt to clarify in more detail the context and the conversation of the different types of treatments being discussed. This has been supplemented by additional healthcare intervention approaches to improve standardisation of documentation of teams transcribing and transferring information relating to ceiling of treatment^5,6^. As a result, there has been an expansion in the vocabulary around advanced directives and end of life care.

Traditional approaches using standardised forms or integrated care pathways to record such sensitive advance care plans have been extremely helpful in recording such complex personalised discussions between healthcare professionals with patients, families and carers^7^. Many of such advance care plans are now captured in standardised electronic form templates often with details captured in typed free-text narrative. Often words and phrases in advance care plans have very specific technical meanings to a specialist which may not match intended meaning as interpreted by a non-specialist or a non-medical individual. Conventionally, studies in this domain have often used qualitative methodologies to disentangle this^8–10^.

To address this quantitative research gap, a computational linguistic approach was used called Natural Language Processing (NLP) to analyse large amounts using unsupervised algorithms to detect patterns in the use of words and phrases, The first approach used a data-driven technique called ‘word2vec’ to represent words from a large body of text in a multi-dimensional vector space (‘latent space’), based on the contextual use of surrounding words^11^. With a sufficient body of text, these ‘word embeddings’ begin to cluster and words that cluster together often have similar meaning. These embeddings therefore follow the philosophical principle first coined by Ludwig Wittgenstein in 1953*”*… *the meaning of a word is its use in the language”*^12^. This ecological data-driven approach has the advantage of also capturing jargon, acronyms and unconventional language that are being used in the real-world.

Using this data-driven approach in a large body of anonymised electronic clinical text at a large urban hospital in London, we analysed whether words or phrases (‘word embeddings’) discussing advance care planning and ceilings of treatment have similar semantic clusters. We also test whether there is any correlation of these ‘word embeddings’ with mortality, and how ‘word embeddings’ are abstracted by AI into ‘concept embeddings’.

## Results

### Word embeddings

The root n-gram “ceiling of treatment” was selected *a priori* by the healthcare team (see Methods), and the 40 n-grams (up to 4 tokens) non-nonsense fragments most closely associated with the root n-gram: “ceiling of treatment” was obtained and shown below. Nonsense fragments included mentions of prescribed drugs like midazolam.

|  |  |
| --- | --- |
| "ceiling of care" | "not for intubation" |
| "ceiling of treatment" | "not for hdu" |
| "ceiling of rx" | "not for icu" |
| "ceilings of care" | "not for itu" |
| "ceilings of treatment" | "not suitable for intubation" |
| "currently for full" | "not suitable for itu" |
| "end of life care" | "palliation" |
| "eol care" | "palliative input" |
| "eolc" | "palliative medications" |
| "escalation beyond" | "palliative treatments only" |
| "limit of care" | "terminal care" |
| "limits of care" | "Treatment Escalation Plan" |
| "not appropriate for itu" | "unsurvivable" |
| "not appropriate for intubation" | "ward based care only" |
| "not for critical care" | "ward based ceiling of care" |
| "not for escalation to itu" | "withdrawal of care" |
| "not for escalation" | "withdrawal of intensive" |
| "not for inotropes" | "withdrawal of treatment" |

The acronyms identified from this approach were easily interpreted to an experienced clinician: “rx” = treatment; “eolc” = end of life care; “itu” = intensive therapy unit; “icu” = intensive care unit; “hdu” = high dependency unit.

### Relationship with outcome

The n-grams above were then divided along phrases with similar meaning (poicelonym), and then repeated word/phrase searches were performed in the whole 2019 inpatient dataset at Kings College Hospital to provide aggregated unique patients with those phrases. There were two broad groups of phrases with similar meaning – phrases relating to the “ceiling of treatment” clusters and “End Of Life”. This is summarised in Table 1 together with proportions with recorded dates of death.

**Table 1:** Word and phrase counts per inpatient were searched across all inpatient records along groups of similar semantics and linked to whether there was an associated date of death. Absolute and relative risk are derived from these absolute values.

|  | Key phrases showing up in documents from Oct-2018 to Sept-2019 | Any inpatients with the phrase during time period in Health Record | Any inpatients with the phrase and Death Dates within 7 days | % of inpatients with phrase and death within 7 days | Relative risk vs annual control |
| --- | --- | --- | --- | --- | --- |
| Ceiling of Care Group | "Treatment Escalation Plan" | 3,181 | 55 | 1.7% | 2.16 |
|  | "not for inotropes" | 20 | <10 | 15.0% | 18.71 |
|  | "not for <u>hdu</u>" | 39 | <10 | 17.9% | 22.33 |
|  | "currently for full" | 83 | 17 | 20.5% | 25.55 |
|  | "ceiling of <u>rx</u>" OR "ceiling of care" OR "ceilings of care" OR "ceilings of treatment" OR "ceiling of treatment" | 1,254 | 203 | 16.2% | 20.20 |
|  | "ceiling of care" OR "ceilings of care" OR "limit of care" OR "limits of care" | 910 | 169 | 18.6% | 23.17 |
|  | "ceilings of treatment" OR "ceilings of treatment" OR "ceiling of <u>rx</u>" | 431 | 54 | 12.5% | 15.63 |
|  | "not for intubation" OR "not suitable for intubation" OR "not appropriate for intubation" | 184 | 51 | 27.7% | 34.58 |
|  | "not for <u>itu</u>" OR "not for <u>icu</u>" OR "not suitable for <u>itu</u>" OR "not appropriate for <u>itu</u>" OR "not for escalation to <u>itu</u>" OR "not for critical care" | 284 | 99 | 34.9% | 43.49 |
|  | "ward based ceiling of care" OR "ward based care only" | 140 | 53 | 37.9% | 47.23 |
|  | "not for escalation" OR "escalation beyond" | 193 | 75 | 38.9% | 48.48 |
|  | "unsurvivable" | 59 | 34 | 57.6% | 71.89 |
| End of Life Care Group | "palliative treatments only" OR "palliative input" OR "palliative medications" OR "palliation" | 1165 | 390 | 33.5% | 41.76 |
|  | "withdrawal of care" OR "withdrawal of treatment" OR "withdrawal of intensive" | 67 | 38 | 56.7% | 70.75 |
|  | "terminal care" OR "end of life care" OR "eol care" OR <u>eolc</u> | 2138 | 1230 | 57.5% | 71.77 |
| Control | None of the above phrases in either cluster | 424905 | 3406 | 0.8% |  |

|  | Key phrases showing up in documents from Oct-2018 to Sept-2019 | Any inpatients with the phrase during time period in Health Record | Any inpatients with the phrase and Death Dates within 7 days | % of inpatients with phrase and death within 7 days | Relative risk vs annual control |
| --- | --- | --- | --- | --- | --- |
| Ceiling of Care Group | "Treatment Escalation Plan" | 3,181 | 55 | 1.7% | 2.16 |
|  | "not for inotropes" | 20 | <10 | 15.0% | 18.71 |
|  | "not for hdu" | 39 | <10 | 17.9% | 22.33 |
|  | "currently for full" | 83 | 17 | 20.5% | 25.55 |
|  | "ceiling of rx" OR "ceiling of care" OR "ceilings of care" OR "ceilings of treatment" OR "ceiling of treatment" | 1,254 | 203 | 16.2% | 20.20 |
|  | "ceiling of care" OR "ceilings of care" OR "limit of care" OR "limits of care" | 910 | 169 | 18.6% | 23.17 |
|  | "ceilings of treatment" OR "ceilings of treatment" OR "ceiling of rx" | 431 | 54 | 12.5% | 15.63 |
|  | "not for intubation" OR "not suitable for intubation" OR "not appropriate for intubation" | 184 | 51 | 27.7% | 34.58 |
|  | "not for itu" OR "not for icu" OR "not suitable for itu" OR "not appropriate for itu" OR "not for escalation to itu" OR "not for critical care" | 284 | 99 | 34.9% | 43.49 |
|  | "ward based ceiling of care" OR "ward based care only" | 140 | 53 | 37.9% | 47.23 |
|  | "not for escalation" OR "escalation beyond" | 193 | 75 | 38.9% | 48.48 |
|  | "unsurvivable" | 59 | 34 | 57.6% | 71.89 |
| End of Life Care Group | "palliative treatments only" OR "palliative input" OR "palliative medications" OR "palliation" | 1165 | 390 | 33.5% | 41.76 |
|  | "withdrawal of care" OR "withdrawal of treatment" OR "withdrawal of intensive" | 67 | 38 | 56.7% | 70.75 |
|  | "terminal care" OR "end of life care" OR "eol care" OR eolc | 2138 | 1230 | 57.5% | 71.77 |
| Control | None of the above phrases in either cluster | 424905 | 3406 | 0.8% |  |

Phrases indicating “End of Life” and “Terminal” clearly had higher rates of mortality since it is implicit in their meaning, whereas terms referring to different limitations of LST had more intermediate prognosis. It is noteworthy that the preferred hospital protocol term to describe such discussions and plans in the hospital -“Treatment Escalation Plan” was extremely common (>3k inpatients). However, this appeared to be used as a heading phrase, as it did not contain any semantic meaning on what the level of advance care was agreed. As a result, the 7-day mortality with “Treatment Escalation Plan” was extremely low. This suggests that these discussions are not foregone conclusions and that having such discussions does not carry an implicit implication of early mortality.

### Concept embeddings

To correct for any mis-spellings and typographical errors, the word embeddings were converted to MedCAT concept embeddings and trained against the entire corpus. To visualise the semantic relationships between these concept embeddings, a t-distributed stochastic neighbour embedding (t-SNE) was used to reduce a high-dimensional vector (300 dimensions) into a 2 dimensional in Figure 1^13^. There are four broad groups which only partially follow the clinical groupings used in Table 1. Of note, the regions outlined in green and red in this 2-dimensional semantic space in Figure 1 correspond to the ‘End of Life’ grouping in Table 1 where the outcomes are poorest. Less discrete clusters in the Blue regions with n-grams of overlapping outcomes describing limits of appropriate interventions similar in meaning to the Ceiling of Care group.

**Figure 1.**
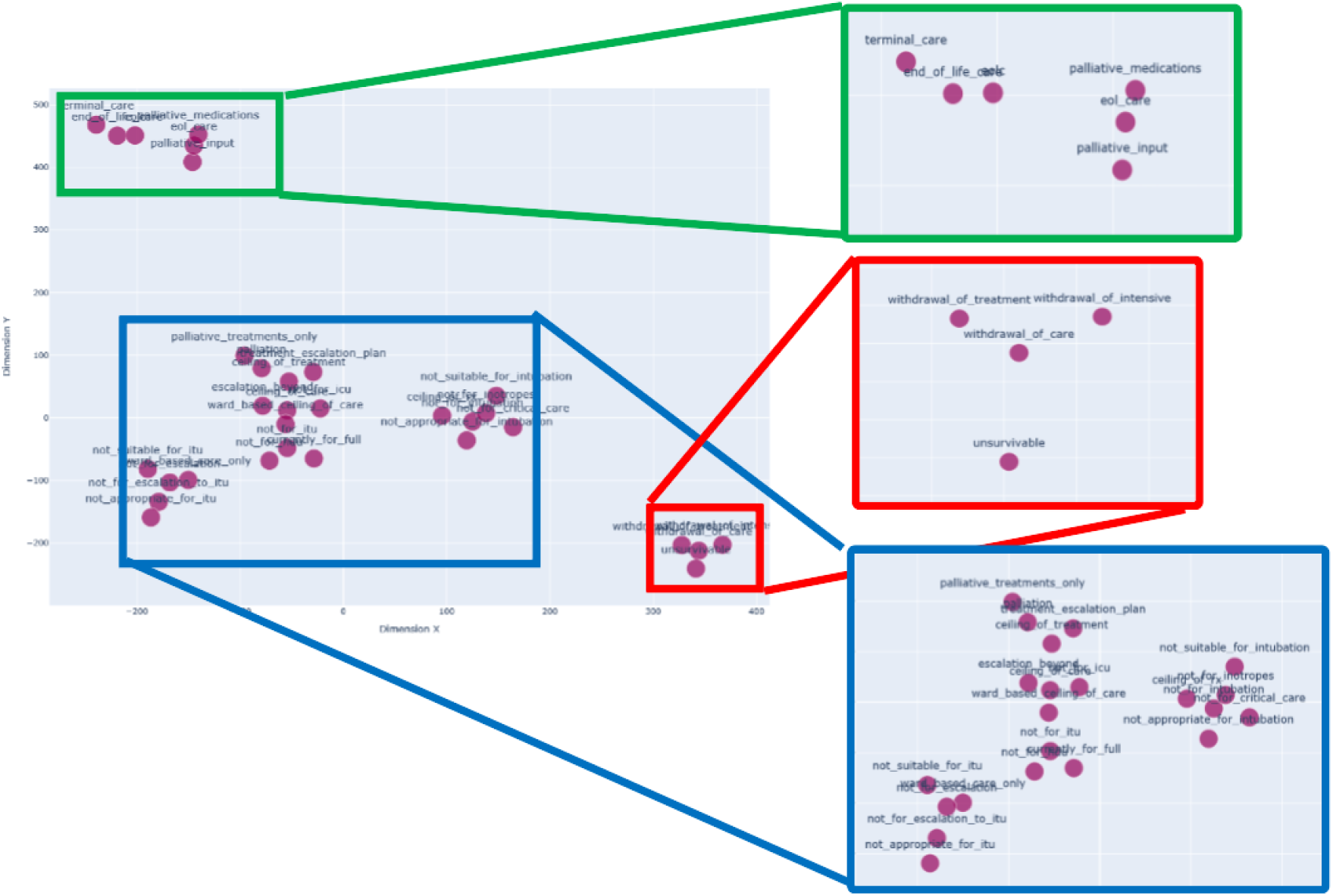
showing the clusters of concept embeddings on a t-distributed stochastic neighbour embedding (TSNE) plot in two-dimensions (X and Y). X and Y represent synthetic dimensions derived from the word embeddings, and is analogous to principle components in a Principal Component Analysis (PCA). Regions of clustering are expanded for clarity with green-red clusters corresponding most similarly to End Of Life Care while blue cluster corresponding to Ceiling of Care. TSNE plot is available as dynamic figure in Supplementary HTML file.

## Discussion

We present the first quantitative NLP evaluation of the language used in real world discussions about ceiling of treatments and End Of Life care. Phrases recording clinical care from the treating medical team had substantial varied language describing advance care planning ranging from specific interventions to terminal prognostication. This study also showed that unsupervised word-embedding techniques (Word2Vec and MedCAT) were able to produce clusters of phrases which reflect phrases of similar meaning using dimensionality reduction techniques. With a sufficiently large corpus of data, such unsupervised NLP techniques were able to capture implicit and inferred poor prognosis.

This study therefore has an inverted design to a previous Sentiment Analysis study of nursing notes from the MIMIC-III public ICU dataset which found a relationship between such ‘sentiment’ with survival^14^; the ‘sentiment’ was calculated using a rules-based semantic analysis tool (TextBlob^15^) designed for generic non-clinical text which assigns a positive or negative ‘sentiment’ score to a piece of text based on the adjectives, verbs and adverbs used in the text^16,17^. In the current study, both an *a priori* approach and an unsupervised clustering approach were used showing clear associations with the ‘ground truth’ of mortality. The derivation of ‘sentiment’ on prognosis from real world clinical text also makes this much more ecological rather than using rule-based text analysis designed for non-clinical uses.

One significant limitation is that this study did not explore temporal trends in prognosis or embeddings. The scope of this study was the ceiling of treatments towards the end of life and so the focus was very much on the discussions and words used very near the end of life (i.e. within the next 7 days). This narrows the vocabulary for prognosis without introducing noise around the vocabulary of tenses and accuracy of time-course prognostication. Another limitation is the lack of distinction between the different types of ceiling of treatment scenarios; it is likely a ceiling of treatment discussion about an elderly disabled patient is substantially different to that of a young patient with a terminal illness or a sudden traumatic event. Both aspects could be improved on with an expanding corpus as well as exploring the temporal relationship with medical and palliative interventions.

During this study, typographical errors and metonymic variations on free text data entry was frequently detected, requiring an addition of a concept embedding approach. These variations in typing suggest clinicians do not simply copy-and-paste templated thoughts for a very ill patient but instead provide contextualised care to the individual (with manually composed typing) even in an era of increasing standardisation of care pathways.

In summary, this is the first real-world NLP study of ‘End Of Life’ care, mapping out how clinical language is used to describe ‘End Of Life’ discussions as well as to produce syntactic phrase or word clusters that capture information on prognosis. This study introduces quantitative NLP techniques into a field which has traditionally used qualitative approaches. Future work could explore the use of language in different professional groups or explore the temporality of interventions before and after such discussions.

## Methods

### Governance

The project operated under London South East Research Ethics Committee (reference 18/LO/2048) approval granted to the King’s Electronic Records Research Interface (KERRI); specific work on end-of-life care research was reviewed with expert patient input on a virtual committee with Caldicott Guardian oversight. Patient and public engagement was sought throughout this project with expert patients approving the projects as well as writing this article.

### Unsupervised Word and Concept Embeddings

The corpus of records consisted of ∼13 million out of a total of 18 million documents over ∼20 years (2001 to 2020) in the Cogstack platform at Kings College Hospital^18^ which pooled data from the structured and unstructured components of the electronic health record using the CogStack ecosystem tools such as DrugPipeline^19^, MedCAT^20^ and MedCATTrainer^21^. This includes all inpatient and outpatient document text with the exclusion of form checklists and scanned documents of insufficient legibility (∼5 million).

The text was first split into words, then put through a phraser which merged separate tokens into phrases before being passed through *word2vec11*. 2,3,4-Grams were calculated from the text using MedCAT which internally relies on gensim^22^. This allowed us to get embeddings for phrases and not just single words. Given a root n-gram (“Ceiling of Treatment”), the most similar n-grams based on the cosine distance between their vector embeddings were collected and again clustered. This resulted in 32 unique phrases.

### Explanation of grouping of Concept-Embeddings into meaning groups

After the top concepts were identified, these tokens were presented to the 3 healthcare professionals (one critical care physician, one palliative care physician and one neurologist) to group into phrases of similar meaning. These phrase groupings were: “ceiling of care” clusters and “end of life”.

A Cogstack elastic query was then performed for phrases within these clusters to generate total aggregate counts of unique inpatients with documents created in 2019 containing these phrases. The creation dates of the documents were also recorded and matched to whether a date of death was also recorded within 7 days of the date of the recorded phrase. Seven days was chosen to limit the analyses to short-term prognostication. Dates of death were recorded based on the inpatient certification of death by doctor. As a control, all documents in the same time period without these phrases were used. The short time-window provides confidence on accuracy on mortality data as any under-counting of outpatient mortality would not significantly impact the data.

### Visualisation of Concept Embeddings

All selected phrases were also converted into MedCAT concepts and the unsupervised training for concepts was again rerun on a dataset consisting of ∼13M documents. This is to avoid problems with spelling mistakes and metonyms and slight variations in the phrasing.

To visualise the relationship between the chosen concepts, t-distributed stochastic neighbour embedding (t-SNE) was used to reduce a high-dimensional vector (300 dimensions) into a 2 dimensional space^13^. In summary, t-SNE converts similarities between data points to joint probabilities and tries to minimize the Kullback-Leibler divergence between the joint probabilities of the low-dimensional embedding and the high-dimensional data. This plot ensures that word embeddings that are close in the high-dimensional space remain close in low-dimensional representation. An alternative dimensional reduction technique (Uniform Manifold Approximation and Projection, UMAP^23^) was also tested and is available as a Supplementary File.

## Data Availability

The data are not publicly available as the source data analysed is unstructured textual data, which carries risk of patient re-identification.

## Acknowledgements

We would like to thank the Kings Electronic Records Research Interface (KERRI), the Cicely Saunders Institute, the NIHR Applied Research Centre South London and the NIHR Maudsley Biomedical Research Centre.

## Author Contributions

Study Design: JTT, PH

Data Collection: JT, ZK, MAA

Data Analysis: JT, ZK

Manuscript Drafting: SL, WG, AQ, HP, SC, SL

Manuscript Criticism: RD, KS, VM, AQ, HP, SC

